# COVID-19 Fatality Rate Classification using Synthetic Minority Oversampling Technique (SMOTE) for Imbalance Class

**DOI:** 10.1101/2021.05.20.21257539

**Authors:** Timothy Oladunni, Justin Stephan, Lala Aicha Coulibaly

## Abstract

SARS-Cov-2 is not to be introduced anymore. The global pandemic that originated more than a year ago in Wuhan, China has claimed thousands of lives. Since the arrival of this plague, face mask has become part of our dressing code. The focus of this study is to design, develop and evaluate a COVID-19 fatality rate classifier at the county level. The proposed model predicts fatality rate as low, moderate, or high. This will help government and decision makers to improve mitigation strategy and provide measures to reduce the spread of the disease. Tourists and travelers will also find the work useful in planning of trips. Dataset used in the experiment contained imbalanced fatality levels. Therefore, class imbalance was offset using SMOTE. Evaluation of the proposed model was based on precision, F1 score, accuracy, and ROC curve. Five learning algorithms were trained and evaluated. Experimental results showed the Bagging model has the best performance.

## I. Introduction

The world has witnessed pandemics such as Ebola [1], Spanish flu [2], yellow fever [3], HIV-AIDS [4], smallpox [5] etc. Towards the end of 2019, the world witnessed another pandemic that has infected millions of people. Thousands of lives have been lost. As of today, May 11, 2021, the United States tops coronavirus cases, follow closely by India and Brazil with records of 32 751 021, 22 992 517 and 15 209 990, respectively. On mortality count, United States, Brazil, and India recorded 582 355, 423 229 and 249 992, respectively [6]. There is no consensus about the origin of this pandemic, however, most scientists believe that the virus originated from Wuhan China and was transferred from animals to humans [7]. It later found its way from China into other parts of the world.

For more than a year, COVID-19 has interrupted civilization. Humanity was forced to face the reality of a new plague. Before the introduction of vaccines, various governments and decision makers introduced different measures and policies to reduce and combat the spread of the disease: Mask mandate was enforced. Social distancing was imposed. Washing of hands was encouraged. Large gathering was outlawed. Some businesses were forced to close. Travel ban was imposed.

Apart from the government regulations, the fear of COVID-19 affected our daily lifestyles: Airports were deserted because planes refused to fly. Online learning became the new normal for students. Conference meetings were moved online. Religious worship became video conferencing. Political rallies were not welcomed. Parks and recreation centers were deserted. No one was not spared the wrath of COVID-19.

Scientists have carried out different studies in combatting the spread of the disease and improve mitigation strategies. Some of the studies involved the exploration of artificial intelligence. For example, the capability of machine learning algorithms to learn from dataset for pattern recognition and knowledge discovery was employed. In [8], the authors aimed to predict COVID-19 fatality rate in India using Machine Learning Models. Linear Regression and Polynomial Regression learning algorithms were trained and evaluated. Per the outcome of the study, the polynomial regression model gave more accurate results. Since the study was limited to India, in the future, the authors plan on extending the proposed models to be trained on datasets from other countries.

In another study, authors [9] used big data analytics to track the spread of the coronavirus. Comparison of tools in big data using machine learning algorithm was a part of their study. Machine learning techniques such as Linear Regression, Polynomial Regression and SVM algorithm was employed. Experimental results were compared using python and Tableau for visualization. According to the study, Linear Regression produced the most accurate results.

Meng et.al [10] used dataset of 366 critical Covid 19 cases including patients who died and those who survived within 14 days after being labelled as high-risk patients. Convolutional neural network was trained and evaluated to predict the probability of covid-19 patients belongings to high-risk and low-risk group. Area under the curve and other evaluation techniques were used to evaluate their models. Evaluation result demonstrated that the proposed model performed well in predicting high risk patient.

While there are different studies on COVID-19 prediction, there seems to be a paucity in machine learning literatures on the algorithmic classification of fatality rate at the county level; specifically, to low, moderate, or high fatality rate. Therefore, the focus of this study is to design, develop and evaluate a county level classification of COVID-19 fatality rate using learning algorithms. The proposed model will classify fatality rate at the county level into low, moderate, or high.

We believe that all stakeholders should have an answer to the question; what is the fatality rate class of this county? A satisfactory answer will help decision makers in crafting policies on effective mitigation strategies to combat and reduce the spread of the disease. It will also help in the prioritization of vaccine to the most affected counties. Travelers and tourists would have an idea about a COVID-19 implication of a county they want to visit. It will also help residents of each county to know the level of COVID-19 in their county.

### This paper is organized as follow

Section II describes the methodology for the study. Section III evaluates the result of our experiments, while section IV compares the results. Sections V, VI and VII discuss the conclusion, implication of the study and future work, respectively. We highlighted the limitation of the study in section VII and acknowledged our funding source in section IX.

## II. Methodology

### 1) Data

Dataset was collected from the John Hopkins University COVID-19 repository [6]. The data comprises of COVID-19 fatality rates information of 3 006 counties of the USA. 171 features were recorded. Information includes city name, state name, fatality rate, age ranges, unemployment rate, race, days etc. Redundant and irrelevant features were removed during the data cleaning phase. We aimed to classify Fatality rate using five learning algorithms. Experimental flowchart is shown in figure 1.

**Figure 1.**
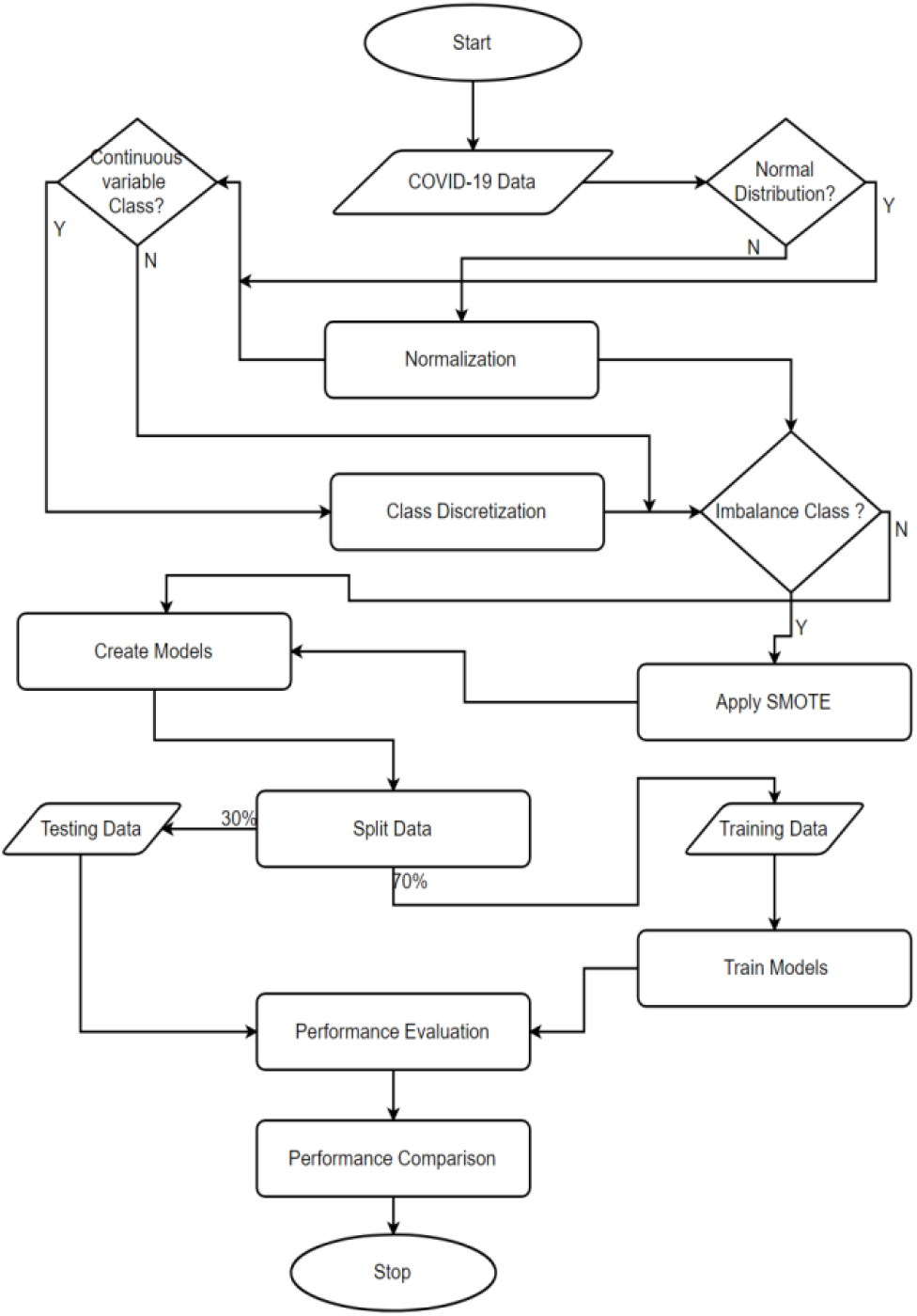
System Flowchart

### 2) Preprocessing

Below is the summary of stages in the preprocessing phase

- Search for null values drop them.
- Drop insignificant and redundant features.
- Discretize.
- Normalize.
- Apply SMOTE.
- Split data into train and test dataset.

#### a) Discretization

Fatality Rate is a continuous variable to be classified into low, moderate, and high fatalities. Therefore, a code was written to convert Fatality Rate to categorical variable for classification purposes. The ‘FatalityRa’ values were converted to classes using the following function:

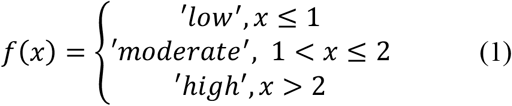

Where *x* is the fatality rate.

Labels were created for each example in the dataset using the ‘FatalityRa’ column. Since this is a classification problem, each of the three labels become the classes: low, moderate, and high.

#### b) Class Imbalance

Imbalance classes are unique cases in machine learning where classes are disproportionally represented [11]. Dividing our datasets into the three classes, Low: (1054,), moderate: (807,), and High: (1281,). The ratio of moderate is much lower than the high and low, suggesting that fatality rate in the USA during the pandemic was at both extremes; either low or high. It has been shown that class imbalance generally produces misleading classification accuracy [12].

#### c) Balancing the Classes

Since the challenge of imbalance class is a major concern in most predictive modelling, some strategies have been developed to reduce its impact. Some of the strategies include 1) collection of more dataset to increase the minority class, 2) change performance metrics for evaluation, 3) apply resampling methodology to the dataset, 4) generate synthetic datasets to augment the minority classes, 5) use ensemble learning algorithms, 6) penalize the models and, 7) apply anomaly detection. Each of these strategies have advantages and disadvantages [13]. For this project, we will explore the generation of synthetic datasets methodology.

#### d) Synthetic Minority Oversampling Technique (SMOTE)

The technique used in this study is the synthetic minority oversampling technique (SMOTE) methodology. SMOTE works by selecting a random sample of the *m* minority class examples. The *k* nearest neighbors of that sample are chosen and a synthetic example is created from a randomly selected point in that region. This is a very effective process to balance imbalanced datasets because it creates approximate examples that are relatively similar to other examples from the minority classes. Thus, the minority classes examples are increased to equalize the dataset.

Scientists and researchers have adopted the use of SMOTE to improve the performance of imbalanced classes. For example, Andrew et.al., implemented SMOTE to improve sentiment analysis of written and spoken language. The proposed model was to predict a person’s expression, emotions, and attitudes [14]. Nuanwan et.al., proposed a Random Forest with SMOTE to improve the predictive capability of online comments on cooking video clips. Per the result of the experiment, the proposed model had a 95% FI score [15].

A SMOTE algorithm has the following sequence:

1. Select a random sample of the *m* minority class examples.
2. Chose the k nearest neighbors of the sample in i.
3. Create synthetic examples from the randomly selected points.
4. Update the minority class with the result of iii.

#### e) Normalization

Dataset was normalized to reduce the impact of large features. It helps to create a level playing field such that each feature has an equal contribution to the model. Dataset are re-scaled to a range of 0 to 1 or -1 to +1. A normalized data improves generalizability [16]. For this study a Min–Max Normalization (MMN) methodology was used. It is mathematically represented as:

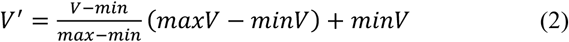

Where the current value V is transformed into the new value V’. maxV and minV are the maximum and minimum values of each column respectively.

### 3) Learning Algortihms

In this study, supervised machine learning approach was used to train and evaluate our models. In supervised learning, labels were shown to the learning algorithm during the training phase. The learning algorithm is expected to learn the mapping of patterns in the datasets from the explanatory variables to the labels. At the testing phase, the learning algorithms are to make predictions based on the learned patterns [17]. We experimented with the following learning algorithms: K-Nearest Neighbor (KNN), Random Forest, Support Vector Machine (SVM), Logistic Regression, Bagging and Boosting. Dataset was divided into 70:30 for training and testing, respectively.

#### a) K-Nearest Neighbor (KNN)

Behind the scene, KNN algorithm calculate distance between training data points and a testing datapoints. The process of classifying elements in a set goes as follows: In a set of elements S, the algorithm find the most similar K training status for a given set. Each status class will be given a value that represents similarity sum between these training elements and the testing elements. The statistical value of each class is obtained with K elements and the scores are sorted.

Assuming k is the nearest number with *n* numbers of example set, KNN follows this steps:

- Step 1: Chose a value of K (initialization)
- Step 2: For *n* training datapoints
  - Step 2.1: Compute the Euclidean distance between the training and testing datapoints
  - Step 2.2: Sort results in ascending order
  - Step 2.3: Choose top K rows
  - Step 2.4: Assign a class to each test row Given two datapoints *x* and *y*, their Euclidean distance is computed as:

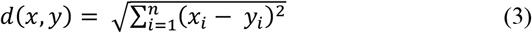

Where *n* is the number of observations.

#### b) Random Forest Algorithm

Random Forest is a supervised machine learning algorithm. It builds forest that are ensemble of decision trees. It generates multiple decision trees by randomly selecting samples and features. The classification results of Random Forest Algorithm of multiple trees are based on the idea that minority is subordinate to the majority. Random forest is most of the time more effective than single trees. An advantage of this algorithm is that it uses a combination of learning models to increase accuracy.

Compared to a single tree, it has been shown that Random forest is more effective in reducing errors [18]. Its ability to decorrelate trees gives it an edge in reducing overfitting and handling of inbalanced classes. It also has the capability of determining features importance. This algorithm can be used for association, regression, as well as classification problem.

Formula: It first perform entropy on the training and testing dataset.

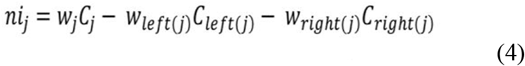

Where,

ni sub(j)= the importance of node j

w sub(j) = weighted number of samples reaching node j

C sub(j)= the impurity value of node j

left(j) = child node from left split on node j

right(j) = child node from right split on node j

#### c) Bagging

This is a decision tree ensemble where repeated samples were taken from the dataset to create and generate multiple training examples. *B* different datasets were trained. We trained *b* bootstrapped models and took the mean of the result. Mathematically:

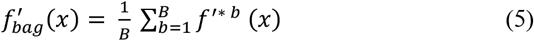

Bagging is an aggregation or ensemble trees for reducing the variance of decision trees. Given *n* predictive variables *Z*_*1*_ to *Z*_*n*_, each with a variance of σ ^2^, the variance of the mean is given as *σ* ^*2*^ */n*. Suggesting an improvement in the predictive capability since the variance is reduced.

#### d) Support Vector Machine Algorithm (SVM)

The SVM is a learning algorithm that improves its predictive capability by enlarging its feature space. Its use of *kernels* enables it to accommodate a non-linear boundary. This arrangement makes it suitable as a non-linear classifier. Given two r vectors *a* and *b*. their inner product is defined as;

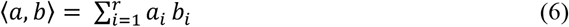

For two of observations *x*_*i*,_ and *xi’*, their inner products is defined as:

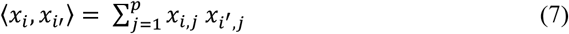

The support vector classifier can be expressed as:

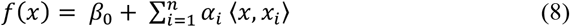

*β*_0_ *and α*_*i*_ are the initial parameter and estimated parameters respectively for *i* to *n* number of training observations.

#### e) Logistic Regression Algorithm

Logistic Regression Algorithm is supervised machine learning algorithm that can be used for classification problem was on probability concept. The algorithm designate observation to discrete values of classes set. It uses the logistic sigmoid function. With logistic sigmoid the cost function only varies between 0 and 1.

The steps for Logistic Regression Algorithm are process are:

- Step 1: Assign value of 0 to each coefficient and calculate probability of the first training instance.
- Step 2: Calculate new coefficient value using update equation.
- Step 3: Repeat the process for all training instance.

Logistic regression can be expressed as

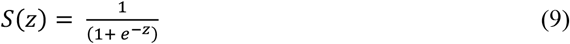

Where:

s(z) = output between 0 and 1

z = input to the function

e = base of natural log

## III. Evaluation

The performance of the learning algorithms was evaluated using class precision, F1 score, accuracy, and ROC curve.

### A) Precision

Precision is defined as the ratio of true positive to true positive plus the false negative. This can be represented mathematically as:

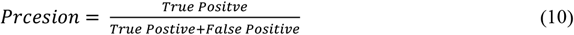

## B) F1 Score

F1 provides s a metric to measure the balance between precision and recall. It is the harmonic mean of the two metrics. This harmonic mean is a constrain on extreme values. Mathematically, F1 score is represented as:

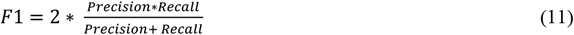

### C) *Accurac*y

Accuracy is the ratio of correct samples predictions to total number of all predictions and shows the general model’s performance in terms of correct classification. It is represented mathematically as:

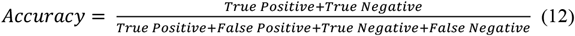

### D) ROC Curve

The performance of the models was visualized using the ROC curve. The ROC curve shows the True Positive Rate (TPR) and False Positive Rate (FPR) plotted on the y-axis and x-axis respectively.

TPR is defined as:

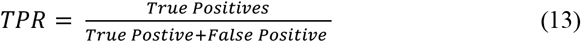

FPR is defined as:

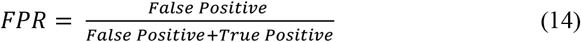

## IV. RESULTS

Using the above metrics, the result of the experiment is shown in tables 1 to 3.

**TABLE 1.**
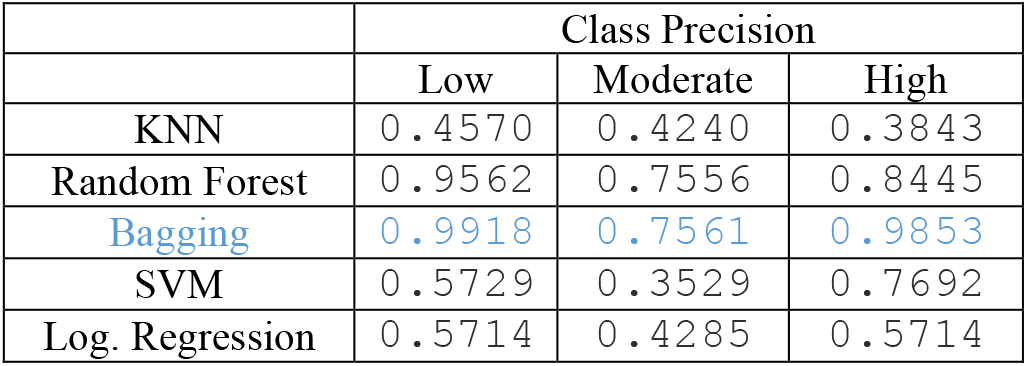
Class Precision

|  | Class Precision |  |  |
| --- | --- | --- | --- |
|  | Low | Moderate | High |
| KNN | 0.4570 | 0.4240 | 0.3843 |
| Random Forest | 0.9562 | 0.7556 | 0.8445 |
| Bagging | 0.9918 | 0.7561 | 0.9853 |
| SVM | 0.5729 | 0.3529 | 0.7692 |
| Log. Regression | 0.5714 | 0.4285 | 0.5714 |

**TABLE 2.** F1-score

|  | Class F1-Score |  |  |
| --- | --- | --- | --- |
|  | Low | Moderate | High |
| KNN | 0.3894 | 0.3598 | 0.3223 |
| Random Forest | 0.8243 | 0.5520 | 0.7308 |
| Bagging | 0.9618 | 0.7369 | 0.9438 |
| SVM | 0.3767 | 0.0299 | 0.0520 |
| Log. Regression | 0.0581 | 0.0153 | 0.0211 |

**TABLE 3.** Accuracy

|  | Avg. Accuracy |
| --- | --- |
| KNN | 0.3103 |
| Random Forest | 0.5963 |
| Bagging | 0.8020 |
| SVM | 0.1054 |
| Log. Regression | 0.0122 |

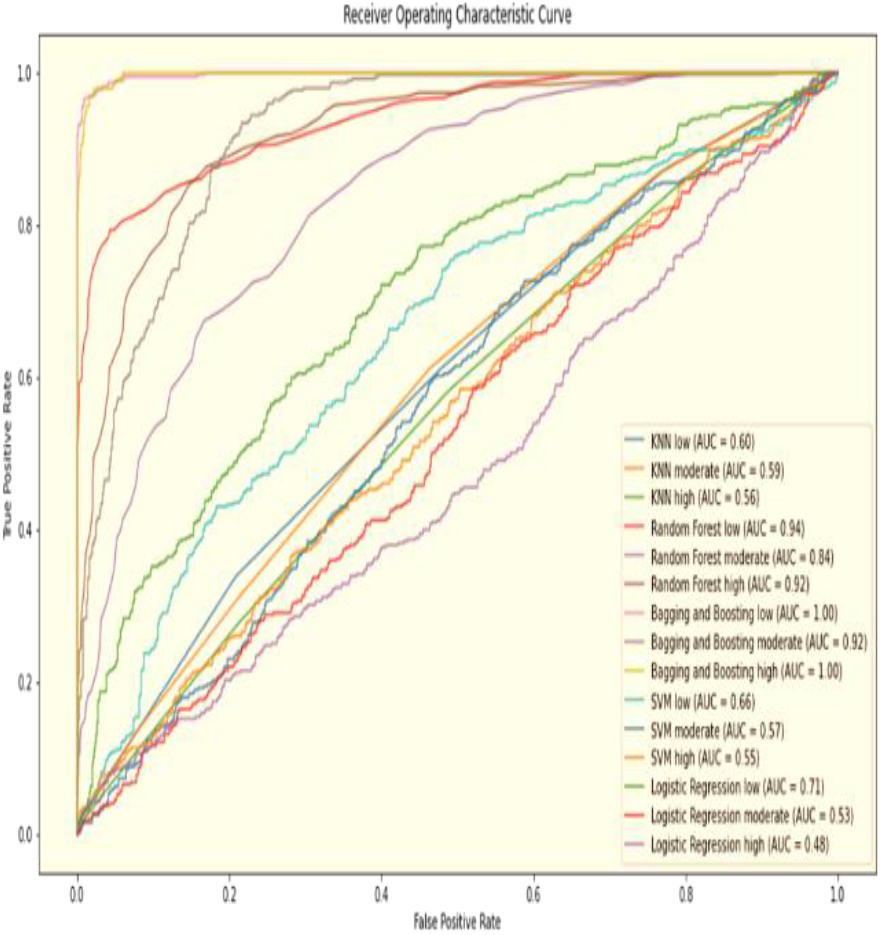

## V. CONCLUSION

COVID-19 pandemic has caused major distraction to the society since its outbreak in 2019. Millions of people have been infected and thousands of lives have been lost. Therefore, in this study, we have designed, developed, and evaluated a COVID-19 fatality rate classifier. Dataset was obtained from the John Hopkins University repository. Classification was based on low, moderate, or high. Imbalanced classes were offset using SMOTE. After addressing the class imbalance problem, the results showed the effectiveness of our strategy.

## VI. Implication of the Study

This study has the following implications:

a. The bagging model has the highest performance based on class precision, F1 score and accuracy.
b. Combining SMOTE with ensemble learning algorithms proved to be the most effective strategy. As shown in our result, Bagging and Random Forest came first and second, respectively.
c. The lowest performing models were SVM and logistic regression with the latter proving to be the worse of the two.
d. Improving the performances of these models may require further adjustments of the hyperparameter values.
e. The high accuracy and precision of the Bagging model suggests that it is has the capability of classifying COVID-19 fatality rates into low, moderate, or high.

## VII. FUTURE WORK

In the future we plan to do the following:

a. Increase classes to 5: very low, low, moderate, high, and very high fatality levels.
b. Experiment with neural networks with the hope of improving the classification accuracy of the model.
c. Apply the proposed model to other epidemiological datasets.

## VIII. LIMITATION OF STUDY

Dataset was based on 3 006 counties in the United States. Therefore, number of counties in other countries may have significant effect on the accuracy of the model.

## Data Availability

Data was obtained from JHU COVID-19 repository

## IX. ACKNOWLDGEMENT

This work is funded by the National Science Foundation grant number 2032345.

